# R3T (Rapid Research Response Team) One-step RT-qPCR kit for COVID-19 diagnostic using in-house enzymes

**DOI:** 10.1101/2020.07.31.20165704

**Authors:** Masateru Takahashi, Muhammad Tehseen, Rahul Salunke, Etsuko Takahashi, Sara Mfarrej, Mohamed A. Sobhy, Fatimah S. Alhamlan, Sharif Hala, Gerardo R. Mandujano, Ahmed A. Al-Qahtani, Fadwa S. Alofi, Afrah Alsomali, Anwar M. Hashem, Asim Khogeer, Naif A. M. Almontashiri, Jae Man Lee, Hiroaki Mon, Kosuke Sakashita, Mo Li, Takahiro Kusakabe, Arnab Pain, Samir M. Hamdan

**Author notes:** **Correspondence**: Samir M. Hamdan. These authors contributed equally: Masateru Takahashi, Muhammad Tehseen.

## Abstract

One-step RT-qPCR is the most widely applied method for COVID-19 diagnostics. Designing in-house one-step RT-qPCR kits is restricted by the patent-rights for the production of enzymes and the lack of information about the components of the commercial kits. Here, we provide a simple, economical, and powerful one-step RT-qPCR kit based on patent-free, specifically-tailored versions of Moloney Murine Leukemia Virus Reverse Transcriptase and *Thermus aquaticus* DNA polymerase termed the R3T (Rapid Research Response Team) One-step RT-qPCR. Our kit was routinely able to reliably detect as low as 10 copies of the synthetic RNAs of the SARS-CoV-2. More importantly, our kit successfully detected COVID-19 in clinical samples of broad viral titers with similar reliability and selectivity as that of the Invitrogen SuperScript™ III Platinum™ One-step RT-qPCR and TaqPath™ 1-Step RT-qPCR kits. Overall, our kit has shown robust performance in both of laboratory settings and the Saudi Ministry of Health-approved testing facility.

## Introduction

In December 2019, an outbreak of a new syndrome characterized by serious symptoms including fever, severe respiratory illness, and acute pneumonia eventually leading to respiratory failure and death was reported in Wuhan city of Hubei province in China. This disease swiftly spread out across the globe resulting in unprecedented preventive measures worldwide ^1-2^. On January 7^th^, 2020, the Chinese health authorities confirmed that this recently discovered syndrome was associated with a new member of the coronavirus (CoV) family closely related to a group of severe acute respiratory syndrome coronaviruses (SARS-CoV) ^3-4^. On February 11^th^, the World Health Organization (WHO) designated the name “coronavirus disease 2019” abbreviated as (COVID-19) to this highly contagious disease and declared it as pandemic (http://www.euro.who.int/en/health-topics/health-emergencies/coronavirus-covid-19/novel-coronavirus-2019-ncov). As of July 2020, COVID-19 resulted in nearly 15.1 million confirmed cases including over 620,000 fatalities globally (https://www.covidvisualizer.com).

The COVID-19 pandemic is caused by the new strain (SARS-CoV-2) classified under genus Betacoronavirus (β-CoV) and subgenus Sarbecovirus ^5-6^. Large numbers of SARS-related coronaviruses (SARSr-CoVs) have been discovered in bats, which are their natural hosts ^7-9^. SARS-CoV-2 is 96 % identical at the whole genome level to bat CoV and shares 79.6 % sequence identity to SARS-CoV ^5^. Coronaviruses are characterized by large, single-stranded (ss), positive-sense RNA genomes ranging from 26 to 32 kilo bases (kb) ^10^. Coronaviruses express their replication and transcription complexes, including RNA-dependent RNA polymerase (RdRp), from a single large open reading frame referred to as ORF1ab ^11^. The viral particle is comprised of four main structural proteins; Spike (S), Membrane (M), Envelope (E), and Nucleocapsid (N) proteins ^12-13^.

A contemporary concern of the COVID-19 pandemic is the need for readily accessible, accurate, efficient and cost-effective diagnostic testing for the detection of SARS-CoV-2 and its associated antibodies in infected individuals. To this end, laboratories, universities, and companies around the world have been racing to develop and produce critically needed test kits. Currently, commercially available COVID-19 detection kits can be broadly divided into viral and antibody tests. The viral test relies on biomolecular assays for the detection of SARS-CoV-2 viral RNA using polymerase chain reaction (PCR)-based techniques or nucleic acid hybridization-related strategies. The antibody test is based on serological and immunological assays that primarily detect antibodies produced by individuals as a result of exposure to the virus. Viral diagnostic tests are quite useful and informative as compared to serological methods, which are feasible only after antibodies have been produced and only provide information about previous infection and not on the current status of the virus in the patient ^14-15^. Therefore, PCR-/nucleic acid hybridization-based techniques including reverse transcriptase loop-mediated isothermal amplification (RT-LAMP) ^16-18^, CRISPR-based assays ^19-21^, Reverse-transcription quantitative PCR (RT-qPCR) ^22^ remain in pratice the most widely applied methods for the detection of RNA viruses. Thanks to its sensitivity, specificity, reliability, and multiplexity, RT-qPCR is currently the gold standard for identifying the presence of SARS-CoV-2 ^5, 23-24^. In the present work-flow at the point-of-care diagnostics, there are two adopted schemes, one-step and two-step RT-qPCR (Figure 1). In the one-step RT-qPCR assay, RT and qPCR reactions can be performed within the same experimental setup. Therefore, the operator can mix all the necessary components at the beginning of the reaction, set the appropriate thermal cycle program in the qPCR machine, and analyse the data. Because of the minimal experimental handling, this platform is the best suited for high-throughput screening environment within a fully automated work-flow. On the other hand, two-step RT-qPCR is a method that combines two separate RT and qPCR reactions. Because each reaction is spatially separated, two-step RT-qPCR gives better control over several experimental parameters; such as the size of RT reaction, *i*.*e*. when the yield of cDNA need to be increased the starting RNA material can be incremented; the adjustability of cDNA template amount, *i*.*e*. flexible amounts of cDNA can be added within the dynamic range of qPCR quantitation; multiple usages of cDNAs, *i*.*e*. multi-genes can be quantified from a single cDNA template. Several commercial companies (Invitrogen, Applied Biosystems, Takara Bio, Bioline, Qiagen, etc.) have developed one-step RT-qPCR kits that can be used in combination with TaqMan probe hydrolysis technology (Applied Biosystems, Foster City, CA). The one-step RT-qPCR kit along with TaqMan probe system is the recommended platform for use by the United States Centers for Disease Control and Prevention (CDC) (http://www.cdc.gov/coronavirus/2019-ncov/lab/rt-pcr-panel-primer-probes.html).

**Figure 1.**
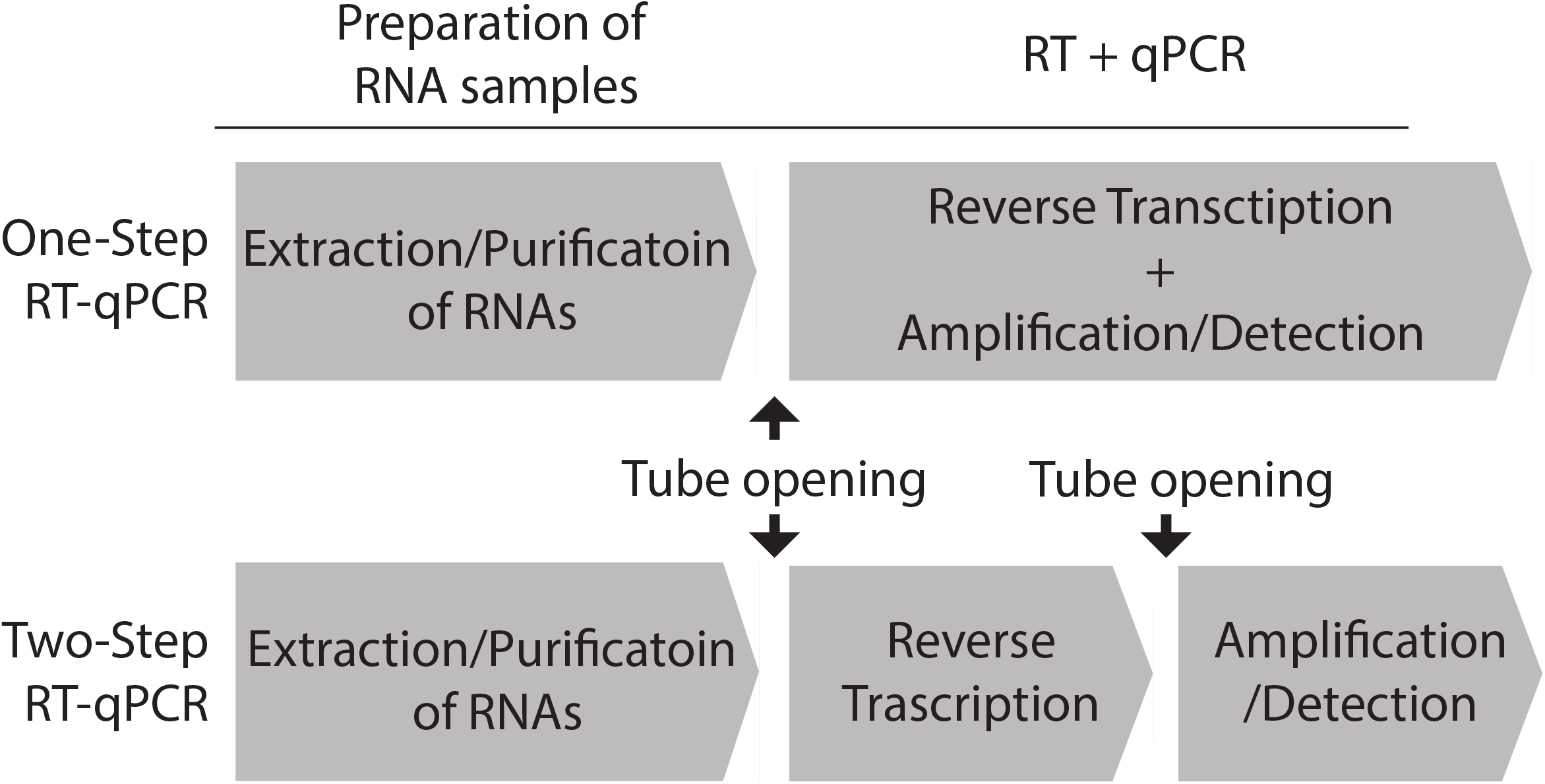
One-step RT-qPCR platform vs. two-step RT-qPCR in the context of work-flow to detect an RNA virus. After sampling the specimen from the patients, RNA materials need to be prepared by extraction and purification (Purification of RNAs). The purified RNAs are converted to DNAs by a reverse transcription reaction using reverse transcriptase (Reverse transcription). At this stage, if the patient was infected with RNA viruses, the complementary DNA (cDNA) fragments derived from the RNA viruses are generated. Then the virus-originated DNA fragments are sufficiently amplified by the qPCR reaction to a detectable level by the fluorescent signals (Amplification/Detection). While the one-step RT-qPCR platform can simultaneously achieve both the RT and qPCR reactions in a single tube, the two-step RT-qPCR needs two separate experimental setups, extra laboratory work, and have more chances for contamination by opening the tubes between the RT and PCR reactions.

Notwithstanding, the facts that one-step RT-qPCR is well-suited for the diagnosis of COVID-19 and that there are many commercially available one-step RT-qPCR kits in the market, their high cost and unavailability due to airport closures and shipment restriction became a major bottleneck that had driven the desire to produce the key components of such kits locally. However, designing in-house one-ste RT-qPCR kits is hampered by the patent-rights for the production of such enzymes and the lack of information about the components of the commercial kits. In this study, we successfully assembled the R3T One-step RT-qPCR kit by purifying two patent-free enzymes; 1) Moloney Murine Leukemia Virus Reverse Transcriptase (MMLV-RT), 2) *Thermus aquaticus* DNA polymerase (Taq Pol). MMLV-RT is a 75 kDa RNA-dependent DNA polymerase that lacks DNA endonuclease activity and has a lower RNase H activity ^25-28^. MMLV-RT is commonly used to synthesize cDNA from ssRNA, ssDNA, or RNA: DNA hybrid templates ^28^. Taq Pol is a 94 kDa DNA-dependent-DNA polymerase that harbours the 5’–3’ but not 3’–5’ exonuclease activity ^29^. The unique 5’–3’ exonuclease activity of Taq Pol cleaves the dual-labeled probe annealed to the target sequence on cDNAs and releases the fluorescent reporter dye, thus recovers its flourecence after it has been quenched by the quencher, making TaqMan probe system ideal reporter in the qPCR reactions.

We used tagged proteins for both Taq Pol (His-Taq Pol) and MMLV-RT (C-His/Strep MMLV-RT) to enhance the expression in *E. coli* and Sf9 cells, respectively, and to make use of the affinitiy chromatography in the protein purification procedures. Purified MMLV-RT is robust enough and supports cDNA synthesis at comparable levels to that of commercially available reverse transcriptases. Our non-hot-start Taq Pol also performs robust amplification in the one-step RT-qPCR platform. By optimizing the reaction buffer besides detemining the optimal amounts of Taq Pol and MMLV-RT used in the one-step RT-qPCR reaction, we circumvent the previously reported inhibition of Taq Pol by reverse transcriptase and therefore sustain the one-step RT-qPCR reaction ^30^. The detection, sensitivity and dynamic range of our R3T One-step RT-qPCR kit (TaqMan probe hydrolysis technology) were evaluated using ten-fold serial dilutions of the standard Twist Synthetic SARS-CoV-2 RNA. The lowest practical detection limit was approximately 10 transcript copies per reaction. In addition, we assessed the performance of our kit using a concise panel of nasopharyngeal swabs of clinical samples from different COVID-19 patients with laboratory confirmed SARS-CoV-2 infection ranging from low to high Ct values. All the samples successfully tested positive with our kit using CDC-verified N1, N2 and RNaseP primer/TaqMan-probe. As a negative control, samples collected during the course of the outbreak from the suspected Saudi patients who were confirmed negative from SARS-CoV-2 infection also tested negative by our kit. Therefore, we conclude here that our R3T One-step RT-qPCR kit can be successfully implemented for routine diagnostics of SARS-CoV-2.

## Materials and Methods

### Expression and purification of Taq Pol

The full-length gene of Taq Pol in pENTR-Taq vector was transferred to our home-made pColdDest vector using Gateway LR reaction (Thermofisher). The resulting plasmid was termed pColdDest-His-Taq (Figure 2B). The expression plasmid of His-Taq Pol was transformed into BL21(DE3) *E. coli* strain, and cells were grown in 10 L LB medium to an OD_600_ of 0.8. The overexpression of His-Taq Pol was induced by 1.0 mM Isopropyl β-D-1-thiogalactopyranoside (IPTG) at 16 °C for 16 hours. The cells were then harvested by centrifugation at 5,500 xg for 15 minutes, re-suspended in Buffer A [50 mM Tris-HCl (pH 7.5), 0.5 M NaCl, 1 mM DTT, 10 % (v/v) Glycerol, 0.5 % NP-40], and incubated on ice with Lysozyme at 1 mg/mL final concentration for 60 minutes. The cells were disrupted by two cycles of sonication (35 % amplitude, 10 second on/off cycle for 5 minutes). Cell debris was removed by centrifugation at 22,040 xg for 30 minutes, and the clear supernatant was collected and incubated at 75 °C for 15 minutes to denature the endogenous proteins from *E. coli*. The heat-denatured solution was then cooled down quickly on ice and centrifuged at 96,000 xg for 45 minutes to remove the denatured proteins. The decanted supernatant was filtered through a 0.45 µM pore size filter and directly loaded onto the His-Trap HP 5 mL (GE Healthcare) column pre-equilibrated with Buffer B [20 mM Tris-HCl (pH 7.5), 0.5 M NaCl] + 20 mM Imidazole. The column was then washed with 20 column volumes (CVs) of Buffer B. The proteins were eluted by 20 CV gradient against Buffer B + 500 mM Imidazole. The peak fractions were analyzed by SDS-PAGE, and the His-Taq Pol containing-fractions were pooled and dialyzed overnight in Buffer C [20 mM Tris-HCl (pH 7.5), 50 mM NaCl, 1 mM EDTA]. The dialyzed sample was then loaded onto the HiTrap SP 5 mL (GE Healthcare) column pre-equilibrated with Buffer C. The proteins were eluted by 20 CV gradient against Buffer D [20 mM Tris-HCl (pH7.5), 1 mM EDTA, 1 M NaCl]. The peak fractions were analyzed by SDS-PAGE and pooled, and the fractions carrying pure His-Taq Pol were dialyzed against Buffer E [50 mM Tris-HCl (pH 8.0), 25 mM NaCl, 0.1 mM EDTA, 1 mM DTT, 0.5 % Tween-20, 0.5 % NP-40, 50 % (v/v) Glycerol]. The dialyzed samples were stored in –20 °C.

**Figure 2.**
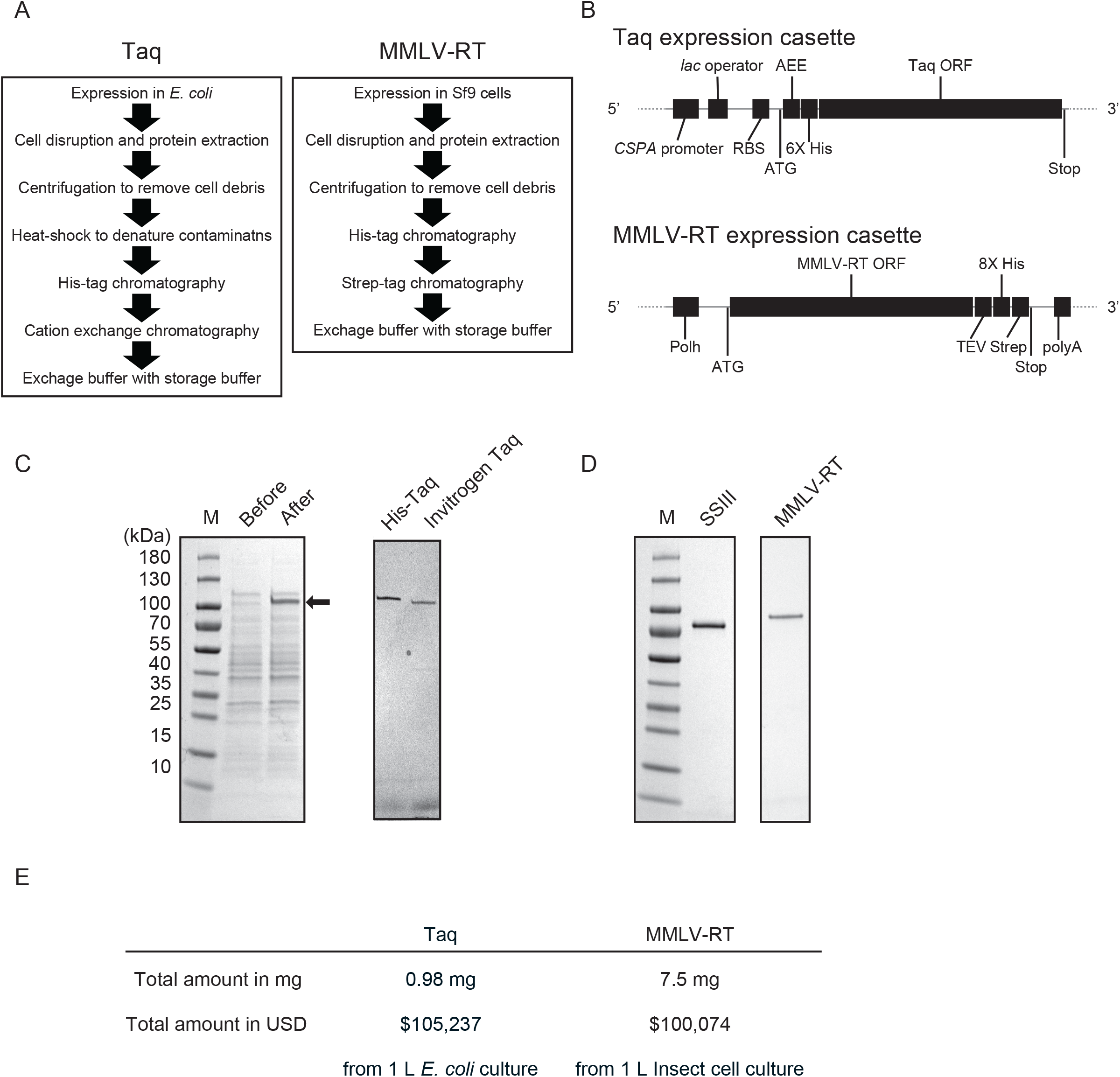
Purification of His-Taq Pol and C-His/Strep MMLV-RT. A) Summary of expression and purification procedures for His-Taq Pol and C-His/Strep MMLV-RT (see the details in the Materials and Methods). B) Schematic representation of the constructs of the recombinant His-Taq Pol and C-His/Strep MMLV-RT expression vectors. CSPA promoter; cold-shock protein A promoter, RBS; ribosome binding site, 6X His; His-tag with 6 histidines, TEE; translational-enhancing element, Taq ORF; Taq Pol open reading frame, Polh; polyhedrin promoter, MMLV-RT ORF; MMLV-RT open reading frame, TEV; Tabacco Etch virus protease targeting site, 8X His; His-tag with 8 histidines, Strep; Strep-tag, polyA; SV40 late polyadenylation signal. C) SDS-PAGE analysis of overexpressed His-Taq Pol in BL21(DE3) *E. coli* cells (left) and purified His-Pol. As a size control, native Taq Pol (N-Taq Pol) (Invitrogen, 18038-018) was applied by 1 µL (5 U). D) SDS-PAGE analysis of purified C-His/Strep MMLV-RT expressed in the Sf9 cells. As a size control, SuperScript III (SSIII, Invitrogen, 18080051) was applied by 1 µL (200 U). E) Summary of closing yields of purified His-Taq Pol and C-His/Strep MMLV-RT.

### Expression and purification of MMLV-RT

The full-length gene of MMLV-RT with cleavable TEV-8xHis-Strep tag at the C-terminus was cloned into the pDEST8 plasmid as previously described ^31^. The plasmid was designated as pDEST8-MMLVRT-TEV-His8-Strept. The C-His/Strep MMLV-RT expression plasmid was then transformed into DH10Bac cells (Life Technologies) to prepare the bacmid DNA for the transfection of Sf9 insect cells. The Sf9 cells were cultured in ESF 921 medium (Expression Systems) at 27°C with continuous shaking at 80 rpm for aeration. To prepare the baculovirus, the C-His/Strep MMLV-RT bacmid DNA was subsequently transfected into Sf9 cells using FuGENE® HD reagent (Promega) per manufacturer’s instructions. The resulting supernatant was collected as P1 virus stock then amplified to obtain P2 virus stock, which was further amplified to generate P3 virus stock for large-scale expression. The expression of C-His/Strep MMLV-RT then proceeded by transfecting 8 L of Sf9 suspension culture at a density of 2 × 10^6^ cells/mL with P3 virus. After 55 hours post transfection with P3 virus, the cells were harvested by centrifugation at 5,500 ×*g* for 10 minutes. The cell pellet was re-suspended in 3 mL per 1 g of wet cells in Buffer F [50 mM Tris-HCl (pH 8.0), 300 mM NaCl, 0.1 % NP-40, 1 mM PMSF, 1 mM EDTA, 5% (v/v) Glycerol, and EDTA-free protease inhibitor cocktail tablets (Roche, UK) at 1 tablet per 50 mL lysis buffer]. All the later steps were performed at 4 °C, where the suspended cells were sonicated, and cell debris was removed by centrifugation at 185,511 xg for 1 hour at 4 °C.

The clear lysate was directly loaded onto the HisTrap Excel 5 mL affinity column (GE Healthcare) pre-equilibrated with Buffer G [50 mM Tris-HCl (pH 8.0), 300 mM NaCl, 0.1 % NP-40, 1 mM EDTA, 5 % (v/v) Glycerol]. The column was then washed with 10 CVs of Buffer G followed by washing with another 10 CVs of Buffer G + 25 mM Imidazole. Finally, the bound proteins were eluted by 10 CV gradient against Buffer G + 500 mM Imidazole. The peak fractions were pooled and dialyzed overnight against Buffer H [100 mM Tris-HCl (pH 8.0), 150 mM NaCl, 1 mM EDTA and 5 % (v/v) Glycerol]. The dialyzed sample was then loaded onto the StrepTrap XT 5 mL column (GE Healthcare) pre-equilibrated with Buffer H. The column was then washed with 10 CVs of Buffer H and eluted by 10 CVs of Buffer H + 50 mM biotin. Fractions containing C-His/Strep MMLV-RT were pooled, dialyzed overnight against storage Buffer I [20 mM Tris-HCl (pH 7.5), 100 mM NaCl, 0.01 % NP-40, 0.1 mM EDTA, 50 % (v/v) Glycerol], then flash frozen in liquid nitrogen, and stored at –80 °C. The protein concentration was determined by measuring the absorbance at 280 nm using an extinction coefficient calculated based on the amino acid sequence of the protein.

### PCR reaction

The PCR reactions by Taq Pol were performed in reaction buffer [20 mM Tris-HCl (pH 8.4), 50 mM KCl, 1.5 mM MgCl_2_, 200 µM dNTPs] besides various amounts of ammonium sulfate and MMLV-RT. pUC19 vector (Invitrogen) was used as a template to amplify the ampicillin resistance gene with the following primers: 5’-TTACCAATGCTTAATCAGTGAGGCACC-3’ and 5’-ATGAGTATTCAACATTTCCGTGTCGCCC-3’. The heating and cooling program used for the PCR reactions involves; pre-denaturation for 2 minutes at 94 °C followed by cycling composed of the three steps: denaturation for 15 seconds, annealing for 15 seconds at 94 °C, and extension for 1 minute at 68 °C. The cycle is then repeated for 29 times.

### Determination of the unit of Taq Pol’s activity

The unit of Taq Pol’s activity was determined based on the standard titeration curve of native Taq Pol’s activity (Thermofisher) (Supplementary Figure 1). Serial dilutions of native Taq Pol (5 U, 2.5 U, 1 U, 0.5 U, and 0.25 U) were used for PCR reactions. The PCR products were analyzed on 1 % agarose gel, and the gel images were captured by iBright Imaging System (Thermofisher). The intensities of the bands corresponding to 862 bp, which is the gene size of ampicillin resistance in pUC19, were quantified by iBright Analysis Software (Thermofisher). The unit of His-Taq Pol’s activity was determined by interpolation from the standard curve using Prism8 software (GraphPad).

### Reverse transcriptase activity assay

The reaction mixture containing 1 µL of RNA template (10^6^ copies/µL of SARS-CoV-2 N gene RNA), 2 µL of random hexamers (3 µM), 1 µL of dNTPs (0.5 mM), and 4 µL of dH_2_O was heated to 65 °C for 5 minutes followed by immediate cooling by placing on ice for more than 1 minute. After the sample tubes were centrifuged briefly, 4 µL of 5x RT buffer, 4 µL of dH_2_O, 2 µL of DTT (10 mM), and 1 µL of RNaseOUT (2 units) (Invitrogen Cat No 10777019) were added, and the mixture was further incubated at room temperature for 2 minutes. Then 1 µL of enzymes was added to the respective reaction of either MMLV-RT, ProtoScript® II (New England Labs, Cat No M0368), NEB AMV (New England Labs, Cat No M0277), Superscript II (Invitrogen, Cat. No 18064022), or Superscript III (Invitrogen, Cat. No 18080093). The final reaction mixtures were incubated at room temperature for 10 minutes, followed by incubation at 42 °C for 50 minutes then by heat inactivation at 70 °C for 15 minutes. Lastly, 5 units of thermostable RNase H (New England Biolabs, Cat. No M0523S) were added and incubated at 37 °C for 20 minutes to hydrolyze RNA. The resulting cDNAs were subjected to RT-PCR assays with 2019-nCoV_N3 primers (IDT, Cat. No 10006770). The RT-PCR reactions were performed on CFX384 Touch Real-Time PCR Detection System (BioRad) using IQ Multiplex Powermix (Bio-Rad, Cat. No 172-5849) and the following program; pre-denaturation at 95 °C for 2 minutes followed by 45 cycles of denaturation at 95 °C for 5 seconds and by annealing/extension at 59 °C for 30 seconds.

### Primer and probe sets

The CDC designed RT-qPCR assay primer and probe set (2019-nCoV CDC EUA kit, Cat. No. 10006606) was purchased from Integrated DNA Technologies (IDT). The kit contains research-use-only primer and probe sets based on the protocol released by CDC (hereafter called CDC assay). The CDC assay includes three sets of the primers and probe labeled with 5’ FAM dye and 3’ Black Hole Quencher® (BHQ).

### DNA/RNA positive controls

The 2019-nCoV-N-Positive Control (IDT, Cat. No. 10006625) is composed of plasmids containing the complete N gene (1,260 base pairs) of SARS-CoV-2. The Hs-RPP30 Positive Control (IDT, Cat. No. 10006626) contains a portion of the ribonuclease P30 subunit (RPP30) gene of the human genome. The control SARS-CoV-2 viral RNA sequences used for constructing RNA titration curves were synthetic RNAs from six sequence variants of the SARS-CoV-2 virus (Twist Bioscience). The dried stock was re-suspended in 100 µL of 1X TE Buffer [10 mM Tris-HCl and 1 mM EDTA (pH 8.0)] to make a stock of 1⨯10^6^ RNA copies/µL.

### Clinical Specimen and RNA extraction

One group of nasopharyngeal swabs (*N = 23*) was collected from COVID-19 suspected patients in Ministry of Health hospitals in the Western region in Kingdom of Saudi Arabia. The swabs were placed in 2 mL screw capped cryotubes containing 1 mL of TRIzol (Ambion) for inactivation and transport to King Abdullah University of Science and Technology (KAUST) for further downstream applications. The sample tubes were sprayed with 70 % ethanol, and RNAs were extracted within 2 hours using the Direct-zol RNA Miniprep kit (Zymo Research) per the manufacturer’s instructions, along with several optimizations to improve the quality and quantity of the extracted RNAs from clinical samples. The optimization also included extending the TRIzol incubation period and the addition of chloroform during the initial lysis step to obtain the aqueous RNA layer. The quality control of purified RNAs was performed using the High Sensitivity Qubit kit (Thermo Fisher) and RNA 6000 Nano Agilent kit (Agilent), respectively.

Another group of nasopharyngeal swabs (*N = 192*) was collected and tested at the Department of Infection and Immunity at King Faisal Specialist Hospital and Research Centre (a Saudi Ministry of Health-approved testing facility). A high-throughput RNA extraction was performed using the RNA KingFisher Flex System and MagMAX Viral/Pathogen Nucleic Acid Isolation Kit (Cat. No. A42352) following the manufacturer’s instructions.

### One-step RT-qPCR

In order to determine the sensitivity of our R3T One-step RT-qPCR kit and detect SARS-CoV-2 in clinical samples, we performed one-step RT-qPCR using primers and probes from the 2019-nCOV CDC EUA Kit produced by IDT (Cat. No. 10006606) or the TaqPath^™^ COVID-19 CE-IVD RT-PCR Kit (Cat. No. A48067). The 2019-nCOV CDC EUA kit contains three sets; each is comprised of two primers and one probe: two sets for the viral nucleocapsid gene (N1 and N2) and one set for the human RNase P gene, according to the guidelines of the CDC diagnostic panel (https://www.fda.gov/media/134922/download). The TaqPath^™^ COVID-19 CE-IVD RT-PCR kit contains TaqPath^™^ COVID-19 Assay Multiplex, which has primers and probes for Gene ORF1ab, N Protein, S Protein, and MS2. Briefly, the reaction mixture of a total 20 μL reaction volume was constituted of 10 μL of 2X reaction buffer mix, 1 μL of His-Taq Pol/C-His/Strep MMLV-RT enzyme mix, 1μL of probe/primer mix, 1 or 2 μL of RNA and nuclease-free water to reach to the final volume. The one-step RT-qPCR was performed in ABI 7500 and 7900 Fast Real-Time PCR system (Applied Biosystems, USA) for the side-by-side comparison with the Invitrogen SuperScript™ III Platinum™ One-step RT-qPCR and Thermofisher TaqPath™ 1-Step RT-qPCR, respectively. The RT-qPCR conditions were as follows: reverse transcription at 55 °C for 30 minutes, pre-denaturation at 94 °C for 2 minutes, followed by cycling composed of the three steps: denaturation at 94 °C for 15 seconds, annealing at 58 °C for 30 seconds, and extension at 68 °C for 1 minute repeated for 45 cycles. At the end, the reaction was heated at 68 °C for 5 minutes.

## Results

### Expression and purification of histidine-tagged Taq polymerase

The expression and purification of the native form of Taq Pol was established in 1991 ^32^. To make both expression and purification processes straightforward, we devised Taq Pol with N-terminal histidine-tag (His-Taq Pol) under the control of the cold-shock promoter. The construct layout of the expression vector for His-Taq Pol is depicted in Figure 2B. The procedures for the expression and purification of His-Taq Pol are outlined in Figure 2A and described in detail in the Materials and Methods section. With our new construct, we can precisely control the induction and expression level by adjusting IPTG concentration and varying the temperature. As shown in Figure 2C, we observed a noticeable overexpression of His-Taq Pol in comparison to the endogenous *E. coli* proteins by incubating at 16 °C with 1 mM IPTG final concentration. In contrast, we did not detect such enhancement for the expression of native Taq Pol (N-Taq Pol) under the control of pET vector system (data not shown). Tagging at the N-terminus with histidines also enabled us to eliminate the time-consuming polyethylenimine precipitation steps in the previous purification protocol ^32^ without affecting the purity of the final product. As shown in Figure 2C, the purity of the His-Taq Pol was comparable to that of the commercially available N-Taq Pol. Owing to the improved expression level of Taq Pol, as illustrated by the final yield of the purified protein from 1 L *E. coli* culture (Figure 2E), we successfully obtained 0.98 mg of pure protein equivalent to ∼$105,000 in market value.

### Expression and purification of double His- and Strep-tagged MMLV-RT

Next, we expressed a C-terminal double His- and Strep-tagged MMLV-RT (C-His/Streo MMLV-RT) protein in insect cells (Sf9) using the baculovirus expression system (Figure 2B). The expression and purification protocols are outlined in Figure 2A and described in detail in the Materials and Methods section. Previous study demonstrated that C-His/Strep MMLV-RT exhibits higher activity compared to the N-terminal tagged one (N-His/Strep MMLV-RT), where both proteins were produced in the silkworms using the silkworm-baculovirus expression vector system (silkworm-BEVS) ^31^. Therefore, we opted for the expression and purification of the C-His/Strep MMLV-RT from Sf9 cell suspension culture in order to obtain homogenous proteins by implementing the previously established protocol ^31^. The final products were assessed to be more than 95 % pure (Figure 2D). Remarkably, the closing yields of the purified proteins are around 7.5 mg per 1 L insect cell culture, i.e. 7.6 times more than that of His-Taq Pol expressed in *E. coli*. These results suggest that the C-terminus Histidine and Strep double-affinity tagged system enhanced the expression of C-His/Strep MMLV-RT in Sf9 cells. Moreover, two consecutive steps of affinity chromatography resulted in minimal loss of the proteins during the purification. Since both the native form and C-His/Strep MMLV-RT showed the same RT activity ^31^, we omitted the cleavage step of the TEV tag in favor of achieving higher final yield of C-His/Strep MMLV-RT.

### Activity assays of Taq Pol and MMLV-RT

In order to evaluate the activities of both His-Taq Pol and C-His/Strep MMLV-RT, we performed PCR for His-Taq Pol and the reverse transcriptase assay for C-His/Strep MMLV-RT (Figure 3A and 3B, respectively). The relatively high concentration of detergents, i.e. 0.5 % NP-40 and Tween-20, in the storage buffer of Taq Pol hindered the accurate determination of its concentration based on UV absorbance measurement due to the high background. Therefore, we decided to determine the protein amount through measuring its PCR amplification activity relative to commercial N-Taq Pol (Invitrogen) (see Materials and Methods). We determined the optimal dilution factor by the storage buffer for purified His-Taq Pol to be 160 times. The dilution circumvents the inhibitory effect of high concentration of His-Taq Pol on its own PCR amplification activity (Figure 3A). We also constructed a standard titration curve of the activity of N-Taq Pol versus enzyme unit (Supplementary Figure 1), enabling us to define and calculate the unit of our purified His-Taq Pol’s catalytic activity (310 units/µL). We also assessed the activity of our purified C-His/Strep MMLV-RT using two-step RT-qPCR in comparison to various commercially-available reverse transcriptases (Figure 3B). The activity of a fixed amount of 200 units containing approximately 1,000 ng enzyme from each commercial reverse transcriptase preparation was compared to that of variable quantities of C-His/Strep MMLV-RT. We found that the activity of our C-His/Strep MMLV-RT within the range of 250-1,000 ng was almost consistent to SuperScript II, NEB Protoscript II, and NEB AMV-RT. Collectively, we confirmed that our purified His-Taq Pol and C-His/Strep MMLV-RT are competent for the PCR and reverse transcription reactions. From now onwards, we will quantify our purified His-Taq Pol in units and C-His/Strep MMLV-RT in nanograms (ng).

**Figure 3.**
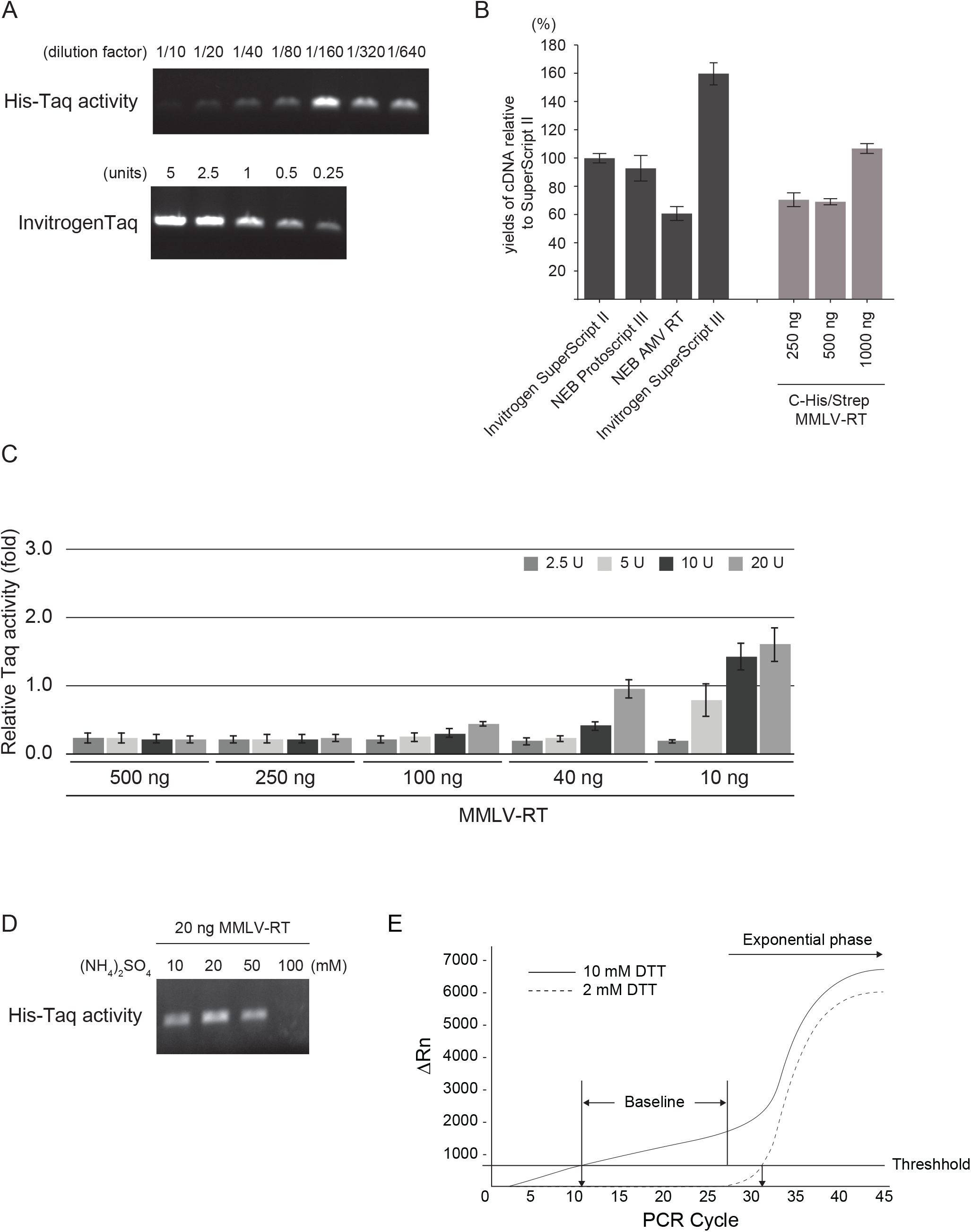
A) Activity assays of His-Taq Pol and N-Taq Pol by PCR. Purified His-Taq Pol was serially diluted by the storage buffer (see Materials and Methods) with the indicated dilution factors and subjected to the PCR reactions. N-Taq Pol is the same as in Figure 2D and the standard titration curve made by N-Taq Pol is shown in Supplemental Figure 1. B) Activity assays of C-His/Strep MMLV-RT and other commercially available reverse transcriptases. The first-strand cDNA was synthesized from SARS-CoV-2 N gene RNAs, and the 2019-nCoV_N3 qPCR assay was conducted. The yields of cDNA synthesis were shown in relative ones compared to that of SuperScript II. Error bars represent standard errors. C) MMLV-RT inhibitory effect on Taq Pol activity in the PCR reaction. The indicated amounts of purified C-His/Strep MMLV-RT were added to the PCR premixture containing different units of Taq Pol. Error bars represent standard errors. D) Ammonium sulfate eases the inhibitory effect of MMLV-RT on the Taq Pol’s activity in PCR. The different amount of ammonium sulfate were added to the PCR reaction mixture without C-His/Strep MMLV-RT, subsequently, 20 ng of C-His/Strep MMLV-RT was added to the reaction. E) The effect of the DTT concentration on the baseline of the TaqMan based detection system. The one-step RT-qPCR reaction was performed with 1,000 copies of the synthetic RNAs as a template. ΔRn; Rn is the fluorescence of the reporter dye divided by the fluorescence of a passive reference dye, ΔRn is Rn minus the baseline.

### Inhibitory effect of MMLV-RT on Taq Pol

To perform two distinct reactions within the same tube, the two enzymes, Taq Pol and MMLV-RT, should work simultaneously, collaboratively, or at least independently without inhibiting one another. However, it was reported by Sellner *et al*. that MMLV-RT inhibits the activity of Taq Pol in the PCR reaction ^30^. To assess the magnitude of the inhibitory effect of MMLV-RT on the Taq Pol’s activity, we performed PCR reactions at varying portions of C-His/Strep MMLV-RT relative to His-Taq Pol (Figure 3C). Per manufacturer’s instructions, the recommended quantity of the reverse transcriptase to synthesize cDNA from total RNAs with dT or random hexamer primers is 200 units (1 µg). Therefore, we started by using 500 ng of C-His/Strep MMLV-RT in combination with the range of 2.5-20 units of His-Taq Pol and gradually decreased the amount of C-His/Strep MMLV-RT in the reaction. We found that C-His/Strep MMLV-RT at 100 ng or more completely inhibits the Taq Pol’s activity even at 20 units of His-Taq Pol. However, by reducing the amount of C-His/Strep MMLV-RT to 40 ng, its inhibitory effect could be overcome by using 20 units of His-Taq Pol. SDS-PAGE analysis of the samples after the reaction showed that the two enzymes remained separate with no apparent cross-linking interaction (Supplementary Figure 2). Furthermore, using size exclusion chromatography (SEC) analysis to evaluate the physical interaction between His-Taq Pol and C-His/Strep MMLV-RT, we could not detect any molecular-size shift in the presence of the two enzymes (Supplementary Figure 3). These results suggest that MMLV-RT hampers the Taq Pol’s activity with a mechanism other than physical interaction. In conclusion, to overcome the inhibitory effect of MMLV-RT on the Taq Pol’s activity, we found that 40 ng MMLV-RT to be the maximum tolerable amount when using 20 units of Taq Pol. Therefore, we decided the mixing ratio to be 2 ng MMLV-RT to 1 unit Taq Pol for subsequent buffer optimization experiments.

### Optimizing buffer components to support one-step RT-qPCR reaction

We also investigated the salt composition of the buffer to further improve the Taq Pol’s activity in the PCR reaction with MMLV-RT. We opted for ammonium sulfate as additional salt and tested different concentrations in the PCR reactions (Figure 3D). We found that the intensity of the PCR product bands gradually increased up to 20 mM of ammonium sulfate followed by decrease at higher concentration of ammonium sulfate due to the suppression of the His-Taq Pol’s activity. Another important parameter for the determination of the optimal buffer composition for the one-step RT-qPCR reaction is DTT concentration. Typically, a relatively high concentration, 10 mM, of DTT is used in the reverse transcription reaction. We investigated the effect of DTT concentration in the context of the one-step RT-qPCR reactions (Figure 3E). At 1 mM DTT, the baseline remained close to zero before the PCR amplification entered into the exponential phase, whereas at 10 mM DTT a gradual increase in the baseline was observed. This baseline drift might be attributed to the degradation of the probe resulting in a higher noise level. With the further optimizations of the one-step RT-qPCR reactions, we reached to the following favorable buffer composition for the subsequent one-step RT-qPCR assays [50 mM Tris-HCl pH (8.5), 75 mM KCl, 2 mM MgCl_2_, 1 mM DTT, 200 uM dNTPs and 13.5 mM ammonium sulfate].

### Determination of the enzyme amounts used in the one-step RT-qPCR reaction, its sensitivity and SARS-CoV-2 detection limit

In order to successfully develop a one-step RT-qPCR platform for the detection of SARS-CoV-2, we formulated and evaluated the efficiency of purified Taq Pol and MMLV-RT in the one-step RT-qPCR scheme using N1 and N2 primer sets and synthetic SARS-CoV-2 RNA as a template. We found that a mixing ratio of 2 ng C-His/Strep MMLV-RT to 1 unit His-Taq Pol still sustained the Taq Pol’s activity in PCR (Figure 3C). Therefore, in our initiail screening, we tried to determine the optimal amount of MMLV-RT in the RT-qPCR reaction while keeping its relative ratio to Taq Pol within this acceptable activity range. Three combinations at diverse His-Taq Pol and C-His/Strep MMLV-RT ratios; 20 U: 40 ng/µL, 30 U: 60 ng/µL, and 40 U: 80 ng/µL were used in each reaction and quantitatively compared to Invitrogen SuperScript™ III Platinum™ One-Step RT-qPCR System (Cat. No. 12574026). We performed three independent replicas of the RT-qPCR assays in triplicates using N1 and N2 primer sets and applying 10-fold serial dilutions of synthetic SARS-CoV-2 RNAs ranging from 10 to 10^5^ copies/µL as a template. The sensitivity of our one-step RT-qPCR system is estimated in terms of the limit of detection (LoD). The LoD assays demonstrated that our one-step RT-qPCR system can reliably detect 40 RNA copies per reaction in all of the three aforementioned combinations (data not shown). Furthermore, our data illustrated that the one-step RT-qPCR assays with 30 U: 60 ng/µL ratio was able to detect as low as 10 RNA copies per reaction from 9 out of 10 replicates as compared to 7 out of 10 replicates in case of 20 U: 40 ng/µL and 3 out of 10 replicates in case of 40 U: 80 ng/µL ratios, indicating the higher sensitivity of our system upon using 30 U: 60 ng/µL ratio. The slope and the R^2^ values of each curve were used to evaluate the efficiency of individual assays (Figure 4). The R^2^ values provided an estimate of the goodness of the linear fit to the data points. In an efficient qPCR assay, R^2^ should be very close or greater than 0.90. The amplification efficiencies of all three His-Taq Pol/C-His/Strep MMLV-RT ratios were above 99% for both N1 and N2 primer sets (Figure 4B, 4C, and 4D). The standard curves showed high correlation coefficients of R^2^>0.99 for N1 primer set and R^2^>0.95 for N2 with the three His-Taq Pol/C-His/Strep MMLV-RT ratios.

**Figure 4.**
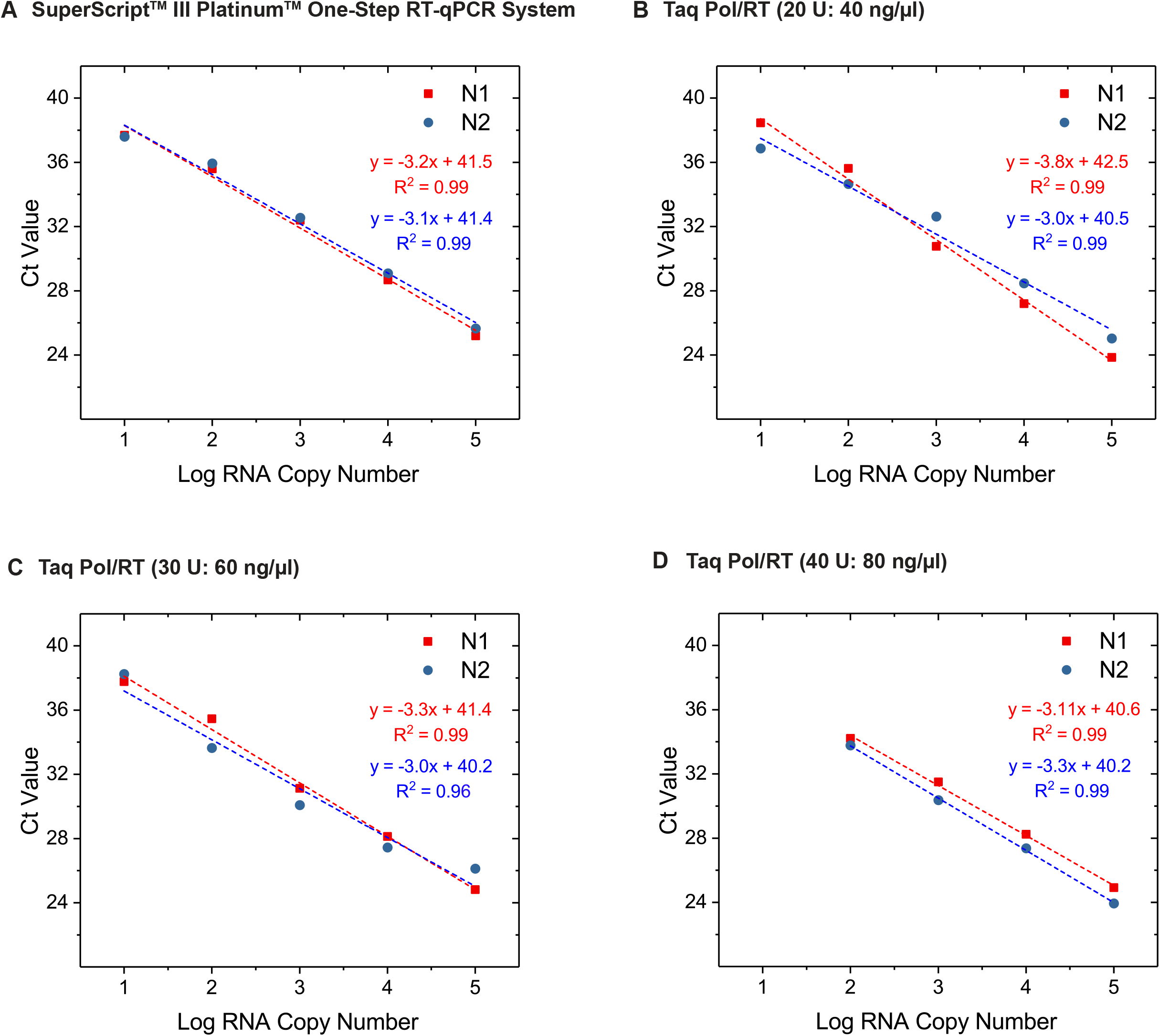
Determination of the optimal proportions of Taq Pol and MMLV-RT in the one-step RT-qPCR reactions. The one-step RT-qPCR reactions were conducted using the synthetic RNAs with 10-fold serial dilutions (from 10 to 10^5^ copies/µL) as a template with N1 or N2 primer sets (See Materials and Methods). As a control, we used (A) SuperScript III Platinum One-Step RT-qPCR kit, (B) 20 units (U) of His-Taq Pol and 40 ng/µL of C-His/Strep MMLV-RT, (C) 30 U and 60 ng/µL, and (D) 40 U and 80 ng/µL. The mixtures were subjected to the reactions and the Ct values were plotted against the threshold cycle. Each plot represents the mean of 3 replicated Ct values with each RNA sample. The coefficient of determination (R^2^) and the equation of the regression curve (y) were calculated and shown in each panel.

### Validation of R3T One-step RT-qPCR system for the detection of SARS-CoV-2 in clinical samples

In order to validate the competency of our one-step RT-qPCR system to detect SARS-CoV-2 in clinical samples from patients, we used RNA samples extracted from the nasopharyngeal swabs of 20 different patients who tested positive besides three patients who tested negative for SARS-CoV-2 using Invitrogen SuperScript™ III Platinum™ One-step RT-qPCR kit (Table 1). The positive samples had variable Ct values ranging from 16 to 38 applying the CDC qPCR *N* gene primer set from IDT. Based on the aformentioned LoD results, we used 30 U: 60 ng/µL ratio for our R3T One-step RT-qPCR kit on these clinical samples. The internal control human RNAseP gene (RP) was detected in all samples. As expected, the N1 and N2 primer pair of the viral N-gene was only detected in the positive samples not the negative ones (Table 1). In case of the three negative control samples (411, 429 and 440), Invitrogen SuperScript™ III Platinum™ One-Step RT-qPCR System gave some high Ct values upon applying N1 primer set but overall result undetermined. The overall Ct values demonstrated a narrow difference between our R3T One-step RT-qPCR kit and Invitrogen SuperScript™ III Platinum™ One-Step RT-qPCR kit (Table 1), demonstrating the sensitivity of our system for the detection of SARS-CoV-2 from clinical samples.

**Table 1:** Comparision of R3T One-stept RT-qPCR System with Invitrogen SuperScript™ III Platinum™ One-Step RT-qPCR System

| R3T one-step RT-qPCR System |  |  |  |  |  |  | Invitrogen SuperScript™ III Platinum™<br>One-Step RT-qPCR System |  |  |  |  |  |  |
| --- | --- | --- | --- | --- | --- | --- | --- | --- | --- | --- | --- | --- | --- |
| Sr. No. | Sample | N1 | N2 | RP | Expected | Result | Sr. No. | Sample | N1 | N2 | RP | Expected | Result |
| 1 | 613 | 16.9 | 17.5 | 27.3 | + | Positive SARS CoV-2 | 1 | 613 | 17.5 | 17.3 | 27.2 | + | Positive SARS CoV-2 |
| 2 | 632 | 17.8 | 18.6 | 27.9 | + | Positive SARS CoV-2 | 2 | 632 | 18.5 | 18.6 | 27.9 | + | Positive SARS CoV-2 |
| 3 | 581 | 20.5 | 21.8 | 25.1 | + | Positive SARS CoV-2 | 3 | 581 | 21.6 | 22.1 | 25.3 | + | Positive SARS CoV-2 |
| 4 | 599 | 22.1 | 23.6 | 23.9 | + | Positive SARS CoV-2 | 4 | 599 | 23.2 | 23.7 | 23.9 | + | Positive SARS CoV-2 |
| 5 | 583 | 24.5 | 25.7 | 25.8 | + | Positive SARS CoV-2 | 5 | 583 | 25.9 | 26.1 | 26.3 | + | Positive SARS CoV-2 |
| 6 | 572 | 27.0 | 28.3 | 24.6 | + | Positive SARS CoV-2 | 6 | 572 | 28.5 | 29.0 | 28.8 | + | Positive SARS CoV-2 |
| 7 | 562 | 30.0 | 30.6 | 28.4 | + | Positive SARS CoV-2 | 7 | 562 | 29.5 | 30.0 | 27.3 | + | Positive SARS CoV-2 |
| 8 | 291 | 26.9 | 29.0 | 23.7 | + | Positive SARS CoV-2 | 8 | 291 | 29.8 | 31.3 | 24.5 | + | Positive SARS CoV-2 |
| 9 | 380 | 27.1 | 27.4 | 23.5 | + | Positive SARS CoV-2 | 9 | 380 | 26.8 | 27.1 | 24.4 | + | Positive SARS CoV-2 |
| 10 | 550 | 32.8 | 33.1 | 26.9 | + | Positive SARS CoV-2 | 10 | 550 | 33.0 | 33.9 | 26.5 | + | Positive SARS CoV-2 |
| 11 | 552 | 32.6 | 35.4 | 30.7 | + | Positive SARS CoV-2 | 11 | 552 | 34.2 | 34.7 | 30.2 | + | Positive SARS CoV-2 |
| 12 | 554 | 33.9 | 35.0 | 31.9 | + | Positive SARS CoV-2 | 12 | 554 | 33.2 | 33.4 | 28.6 | + | Positive SARS CoV-2 |
| 13 | 585 | 35.5 | 38.4 | 24.1 | + | Positive SARS CoV-2 | 13 | 585 | 35.4 | 37.6 | 23.8 | + | Positive SARS CoV-2 |
| 14 | 600 | 35.3 | 38.3 | 26.2 | + | Positive SARS CoV-2 | 14 | 600 | 35.9 | 37.2 | 26.3 | + | Positive SARS CoV-2 |
| 15 | 601 | 34.6 | 36.7 | 23.4 | + | Positive SARS CoV-2 | 15 | 601 | 35.2 | 36.1 | 23.0 | + | Positive SARS CoV-2 |
| 16 | 745 | 34.0 | 37.0 | 28.1 | + | Positive SARS CoV-2 | 16 | 745 | 35.3 | 36.7 | 28.1 | + | Positive SARS CoV-2 |
| 17 | 748 | 31.3 | 33.4 | 27.0 | + | Positive SARS CoV-2 | 17 | 748 | 32.1 | 34.1 | 26.9 | + | Positive SARS CoV-2 |
| 18 | 749 | 34.3 | 36.1 | 32.9 | + | Positive SARS CoV-2 | 18 | 749 | 34.1 | 34.7 | 32.5 | + | Positive SARS CoV-2 |
| 19 | 751 | 35.0 | 37.2 | 29.6 | + | Positive SARS CoV-2 | 19 | 751 | 34.9 | 36.1 | 28.6 | + | Positive SARS CoV-2 |
| 20 | 750 | 34.5 | 38.3 | 37.6 | + | Positive SARS CoV-2 | 20 | 750 | 35.2 | 36.8 | 36.7 | + | Positive SARS CoV-2 |
| 21 | 411 | - | - | 27.5 | - | Negative SARS CoV-2 | 21 | 411 | 36.9 | - | 29.1 | - | Negative SARS CoV-2 |
| 22 | 429 | - | - | 26.9 | - | Negative SARS CoV-2 | 22 | 429 | 39.3 | 40.5 | 27.5 | - | Negative SARS CoV-2 |
| 23 | 440 | - | - | 25.1 | - | Negative SARS CoV-2 | 23 | 440 | 37.6 | - | 25.5 | - | Negative SARS CoV-2 |
| 24 | + | 24.8 | 25.1 | 24.8 | - | Positive Control | 24 | + | 23.6 | 23.6 | 24.2 | - | Positive Control |
| 25 | NTC | - | - | - | - | Not Detecgtd | 25 | NTC | - | - | - | - | Not Detecgtd |

N1 and N2: N-gene; RP: RNaseP; NTC: No template

We next evaluated the performance of our R3T One-step RT-qPCR kit under actual testing conditions in a Saudi Ministry of Health-approved testing facility at King Faisal Specialist Hospital and Research Centre. A total of 192 de-identified patients who were screened for SARS-CoV-2 infection were included in this validation. In a side-by-side comparison of our kit with their endorsed TaqPath™ 1-Step RT-qPCR kit, 20 positive patient samples were identified by both kits (Supplementary Table 1). The remaining 172 patient samples tested negative in both kits; the representative results of these negative patient tests are shown in Supplementary Table 1. This consistency demonstrates the robustness and the accuracy of our optimized R3T One-step RT-qPCR kit’s performance under real testing conditions.

## Discussion

SARS-CoV-2 has originated the present international outbreak of a severe respiratory syndrome termed as COVID-19. The outburst of this viral infection has become a pandemic and severe public health challenge worldwide. Owing to the absence of effective medicines and vaccines, quick identification of the infected individuals and imposing self-quarantine measurements are the only effective containment strategies to avoid widespread community transmission. Therefore, the rapid development of low-cost, easy-to-make, yet sensitive and reliable diagnostic tests are crucial.

In this study, we devised a sensitive, cost-effective, in-house one-step RT-qPCR system based on patent-free Taq Pol and MMLV-RT. These enzymes were discovered in the 1970s ^33-34^; however, they still play a key role as molecular biology tools owing to their robustness and unique enzymatic properties. To avoid legitimate concerns regarding the intellectual property rights, we opted to purify those patent-free enzymes and optimize their use for the one-step RT-qPCR platform. To simplify the expression and purification procedures, we used His-Taq Pol under the control of the cold-shock promoter for *E. coli* expression and C-His/Strep MMLV-RT based on the baculovirus expression system in Sf9 cells. Our protein-tag strategy enabled us to express both proteins in large amounts and apply affinity chromatography for purification without time-consuming precipitation steps. Because our protein purification procedure still includes two chromatography techniques with different chemical properties, the purity of our final product is more than 90 %. Moreover, the high yields of both purified His-Taq Pol and C-His/Strep MMLV-RT proteins from relatively small volumes of 1 L *E. coli* and 1 L Sf9 cultures, respectively, amassed for purified protein amounts of more than $100,000 per enzyme in market value. The quantities of Taq Pol and MMLV-RT produced by our protein expression and purification schemes at the laboratory-scale are satisfactory and do not require production upscaling to an industrial-scale facility. Our protein production platform provides a simpler follow-up example for groups in research institutes with limited-resources.

Till present, there have been many studies for enhancing and improving the enzymatic properties of MMLV-RT and Taq Pol. However, the improvements in the enzymatic performance of commercial MMLV-RTs variants are mainly to amend the efficiency to generate cDNA libraries from total RNAs through random annealing of primers. On the other hand, commercial Taq Pol variants have been improved to enhance the rate and processivity for quicker PCR reactions and longer amplifications. In the scheme of COVID-19 detection, the RT primers are explicitly designed to be gene-specific, besides the RNA regions annealed by the RT primers are carefully selected to avoid erroneous interference in cDNA synthesis. We assume that the well-designed RT primers for cDNA synthesis conceals any possible enzymatic incompetency of our MMLV-RT and enables the detection of COVID-19 RNAs even at low threshold (see discussion below). Our His-Taq Pol is also not a hot-start polymerase like Platinum Taq Pol that we used as a benchmark for performance comparison during this study. However, the advantages of the hot-start PCR polymerase, such as the prevention of primer-dimer formation and non-specific amplification, were not pronounced for the TaqMan based RT-qPCR platform. We also found the use of the relatively short fragments for amplification to be in favor for our His-Taq Pol in the one-step RT-qPCR scheme. We believe that the synergy between using TaqMan based detection system and the short size of amplicons makes the performance of our non-hot-start His-Taq Pol equally robust to that of hot-start Platinum Taq Pol in the one-step RT-qPCR reaction.

Our aim from the current study was to establish the one-step RT-qPCR kit with in-house enzymes. The advantages of a one-step RT-qPCR platform over a two-step process in the work-flow at the point-of-care diagnostics include minimal sample handling, reduced bench time, and less chances for pipetting errors and cross-contamination (Figure 1). This makes the one-step platform the first choice for high-throughput mass screening in the diagnostic point-of-care tests. In the one-step platform, it is crucial to carefully choose the optimal conditions for both MMLV-RT and Taq Pol to work together in the same reaction (1, 2). A major obstacle in assembling our one-step RT-qPCR system was the inhibition of the Taq Pol’s activity in the presence of MMLV-RT. It was proposed that both enzymes interact with specific combination of primers (DNAs) and templates (RNAs) causing inhibitory effect (2). We found that MMLV-RT hinders the Taq Pol’s activity in a portion-dependent manner; however, this inhibitory effect can be circumvented, to some extent, by increasing the amount of Taq Pol. Apart from Taq Pol and MMLV-RT ratio, sulfur-containing inorganic molecules are also known to relieve the inhibition of PCR and often added when using compositions containing two or more enzymes for the reverse transcription activity (US Patent 9,556,466 B2). In this study, besides KCl in the reaction buffer, we employed ammonium sulfate as additional salt and showed that it helped alleviate the MMLV-RT’s inhibitory effect on the Taq Pol’s activity. However, we also noticed that this alleviation disappeared when Taq Pol and MMLV-RT were pre-mixed in the absence of salts (data not shown). This led us to suspect that Taq Pol and MMLV-RT directly interact with each other. However, both the SDS-PAGE and SEC analysis did not show any detectable physical interaction between Taq Pol and MMLV-RT; thus the cause for this adverse effect remains unclear. We strongly believe that the next direction for improving the one-step RT-qPCR, mainly targeted for diagnostic purposes, should focus on finding a way to reduce the MMLV-RT’s inhibitory effect on the Taq Pol’s activity rather than to boost their individual enzymatic activities.

Next, we verified the ideal proportions of Taq Pol and MMLV-RT in the optimized reaction buffer in the context of COVID-19 detection work-flow. We confirmed that 30 U: 60 ng/µL is the optimal composition for our one-step RT-qPCR platform, where we successfully detected as low as 10 RNA copies per reaction with more than 90 % reproducibility. The optimal amounts of MMLV-RT determined here along with the aforementioned ratio for MMLV-RT and Taq Pol are surprisingly low; the manufacturer’s recommendations for making cDNA libraries is 200 units, which is almost 1,000 ng of enzyme for the RT reactions with random hexamer or dT primers. We assume, at least in our case, that lowering the amount of MMLV-RT in the reaction is the only way to make Taq Pol functional in the one-step RT-qPCR platform and that around 60 ng of MMLV-RT is sufficient to synthesize necessary cDNA for COVID-19 diagnostic purposes.

Finally, we assessed our R3T One-step RT-qPCR kit with actual patient samples isolated from individuals infected by SARS-CoV2. We screened 23 different patients under laboratory settings, including three who were suspected but tested negative for SARS-CoV2. All the tests performed with our kit showed the expected true positive and negative results with a slight difference in the Ct values obtained from Invitrogen SuperScript™ III Platinum™ One-Step RT-qPCR. Furthermore, we evaluated the performance of our R3T One-step RT-qPCR kit in a Saudi Ministry of Health-approved testing facility using 192 patient samples who were screened for SARS-CoV-2 infection. Our kit matched their endorsed TaqPath™ 1-Step RT-qPCR kit by identifying the same 20 positive and 172 negative cases, demonstrating that our kit is ready for deployment in the actual testing facilities. Here, we provide a guide on how to assemble an in-house one-step RT-qPCR kit based on patent-free enzymes within a few weeks of conceptualizing the project. We believe that our one-step RT-qPCR system can be successfully applied for the routine SARS-CoV2 diagnostics and extended to detect other pathogens.

## Data Availability

We included all data in the manuscript including Supplementary Figures and Table.

## Acknowledgements

This research was funded by baseline funding from KAUST to S.M.H. and COVID-19 response initiative by the vice president of research at KAUST. The clinical COVID-19 samples were collected as part of KAUST baseline funding (BAS/1/1020-01-01) to AP and the R3T initiative by the the vice president of research at KAUST. The authors declare no conflicts of interest.

## Author Contributions

J. M. Lee, K. Sakashita and H. Mon perfomed molecular cloning of the expression plasmids. M. Takahashi, M. Tehseen, and E. Takahashi perfomed protein purifications and bulk assays. M. Tehseen, R. Salunke, F. S. Alhamlan, and A. A. Al-Qahtani conducted RT-qPCR assays. G. R. Mandujano and M. Li perfomed RT assays. S. Mfarrej and A. Pain extracted RNA samples from the patients. S. Hala, F. S. Alofi, A. Alsomali, A. M. Hashem, A. Khogeer, and N. A. M. Almontashiri collected Nasopharyngeal swabs for the laboratory setting study and A. A. Al-Qahtani and A. Al-Qahtani for the testing facility setting study. This study was initiated by M. Takahashi, T. Kusakabe, and S. M. Hamdan and designed by M. Takashi, M. Tehseen and S. M. Hamdan. M. Takahashi, M. Tehseen, M. A. Sobhy, and S. M. Hamdan wrote the manuscript, and all authors read and commented on the manuscript. M. Takashi, M. Tehseen, A. Pain and S. M. Hamdan supervised the study.

